# Study protocol: Medium throughput, deep proteomic characterization of children with PIMS-TS, and identification of candidate diagnostic biomarkers

**DOI:** 10.1101/2022.12.27.22283890

**Authors:** Cathal Roarty, Clare Mills, Claire Tonry, Peter Cosgrove, Hannah Norman-Bruce, Helen Groves, Chris Watson, Tom Waterfield

**Affiliations:** Wellcome-Wolfson Institute for Experimental Medicine, Queen’s University Belfast- School of Medicine, Dentistry and Biomedical Sciences, Belfast, UK; Paediatric Intesive Care Unit, Royal Belfast Hospital for Sick Children, Belfast, United Kingdom; Padiatric Infectious Diseases,Royal Belfast Hospital for Sick Children, Belfast, United Kingdom

## Abstract

SARS-CoV-2 infection in children results in a wide range of clinical outcomes. Paediatric Multisystem Inflammatory syndrome temporally associated with COVID-19(PIMS-TS) occurs weeks after a SARS-CoV-2 infection, and results in severe illness. This protocol describes a study to fully characterize the circulating proteome of children who have PIMS-TS, the proteome of healthy children who have previously been infected with SARS-CoV-2 and the proteome of febrile children with a confirmed invasive infection. Orthogonal proteomic techniques will be utilized to provide a deep proteomic characterization.

## Background

Paediatric COVID-19, caused by SARS-CoV-2 infection, has a wide variation in clinical outcome, from asymptomatic infection to hospitalization in rarer cases.[1–3] In addition, a small proportion of children infected with SARS-CoV-2 progress to a multisystem hyperinflammatory condition called Paediatric Multisystem Inflammatory syndrome temporally associated with COVID-19(PIMS-TS).[4] This occurs approximately four weeks after initial infection, with most children having antibodies to SARS-CoV-2 and history of an exposure to SARS-CoV-2. There is an estimated one case of PIMS-TS per ten thousand infected children. These children are severely ill, requiring hospital admission and immunomodulatory treatment. The case definitions used by the World Health Organization, the Royal College of Paediatricians and Child Healthy and the Centres for Disease Control and Prevention are in Table 1.[5–7]

**Table 1.** RCPCH, CDC and WHO case definitions for PIMS-TS

|  |  |  |
| --- | --- | --- |
| RCPCH | CDC | WHO |
| A child with persistent fever, inflammation and evidence of organ dysfunction with additional features. | An individual aged <21 years presenting with fever, laboratory evidence of inflammation, with evidence of clinically severe illness requiring hospitalization, with $\geq 2$ organ involvement. | Children and adolescents 0–19 years of age with fever $\geq 3$ days, and two of the following:<br>• Rash, conjunctivitis or muco-cutaneous inflammation signs<br>• Hypotension/sock.<br>• Evidence of myocardial dysfunction, coronary abnormalities or pericarditis<br>• Evidence of coagulopathy<br>• Acute gastrointestinal problems. |
| **SARS-CoV-2 Exposure/Serology** |  |  |
| SARS-CoV-2 PCR testing may be positive or negative | Positive for SARS-CoV-2 infection by RT-PCR, serology, or antigen test.<br>Alternatively exposure to a suspected or confirmed COVID-19 case within the 4 weeks prior to the onset of symptoms. | Evidence of COVID-19 (RT-PCR, antigen test or serology positive), or likely contact with patients with COVID-19. |
| **Exclusion criteria** |  |  |
| Exclusion of any other microbial cause, including bacterial sepsis, staphylococcal or streptococcal shock syndromes, infections associated with myocarditis. | No alternative plausible diagnoses | No other obvious microbial cause of inflammation, including bacterial sepsis, staphylococcal or streptococcal shock syndromes. |

The COVID Warriors study is a prospective multicentre observational cohort study which identified the symptoms associated with SARS-CoV-2 infection in children, and the longitudinal kinetics of anti-SARS-CoV-2 antibodies in children.[1,8,9] With the emergence of PIMS-TS, and it’s temporal link with SARS-CoV-2 infection, an amendment to the study protocol was obtained to allow a cohort of PIMS-TS cases to be recruited from one of the participating sites.

### Rationale

PIMS-TS presents clinicians with several challenges. It was initially reported as an atypical Kawasaki disease presentation, with many shared features such as fever, elevated inflammatory markers, myocarditis and dermatological involvement.[10,11] As has been well documented, it can be a challenging diagnosis, with many case reports of PIMS-TS being mistaken for appendicitis.[12–14] Additionally, there can be a range of severity in PIMS cases, with some children very unwell, requiring ICU admission, usually for inotropic support, while other children can be managed in a ward setting without the need for inotropes. [15]

Misdiagnosing a child with an acute severe viral or bacterial infection with PIMS-TS is a possibility, given the similarities in their initial condition when presenting. It would be clinically useful to have a diagnostic biomarker or panel of biomarkers which can be used to identify children with PIMS-TS. To date, no study has combined the unbiased approach of mass spectrometry with the high sensitivity of proximity extension assay (PEA) technology to provide a deep characterization of the circulating proteome of PIMS-TS. In this study, an unbiased discovery proteomics approach will be employed to achieve this. In addition the comparison of the PIMS-TS proteome with the proteome of febrile and healthy controls will be used to identify prospective candidate diagnostic biomarkers for PIMS-TS.

### Objectives

1. Characterize the circulating plasma proteome of children with PIMS-TS using mass spectrometry and PEA technology.
2. Describe the clinical characteristics and outcomes of children with PIMS-TS.
3. Compare the proteome of children with PIMS-TS to the proteome of febrile children and identify candidate diagnostic biomarkers.
4. Identify the biological roles of differentially abundant proteins identified in PIMS-TS using a literature search, creation of a gene-gene /protein-protein interaction network using CYTOSCAPE or PINE/STRING or other protein network analysis.[16,17]

## Methods

### Study design

A prospective observational matched cohort study characterizing the proteome of children with PIMS-TS and identifying candidate biomarkers by comparing with the proteome of healthy and febrile controls, outlined in figure 1. The study design has been written in conjunction with Recommendations for biomarker identification and qualification in clinical proteomics and the Strengthening the Reporting of Observational Studies in Epidemiology (STROBE) Statement.[18,19]

**Figure 1.**
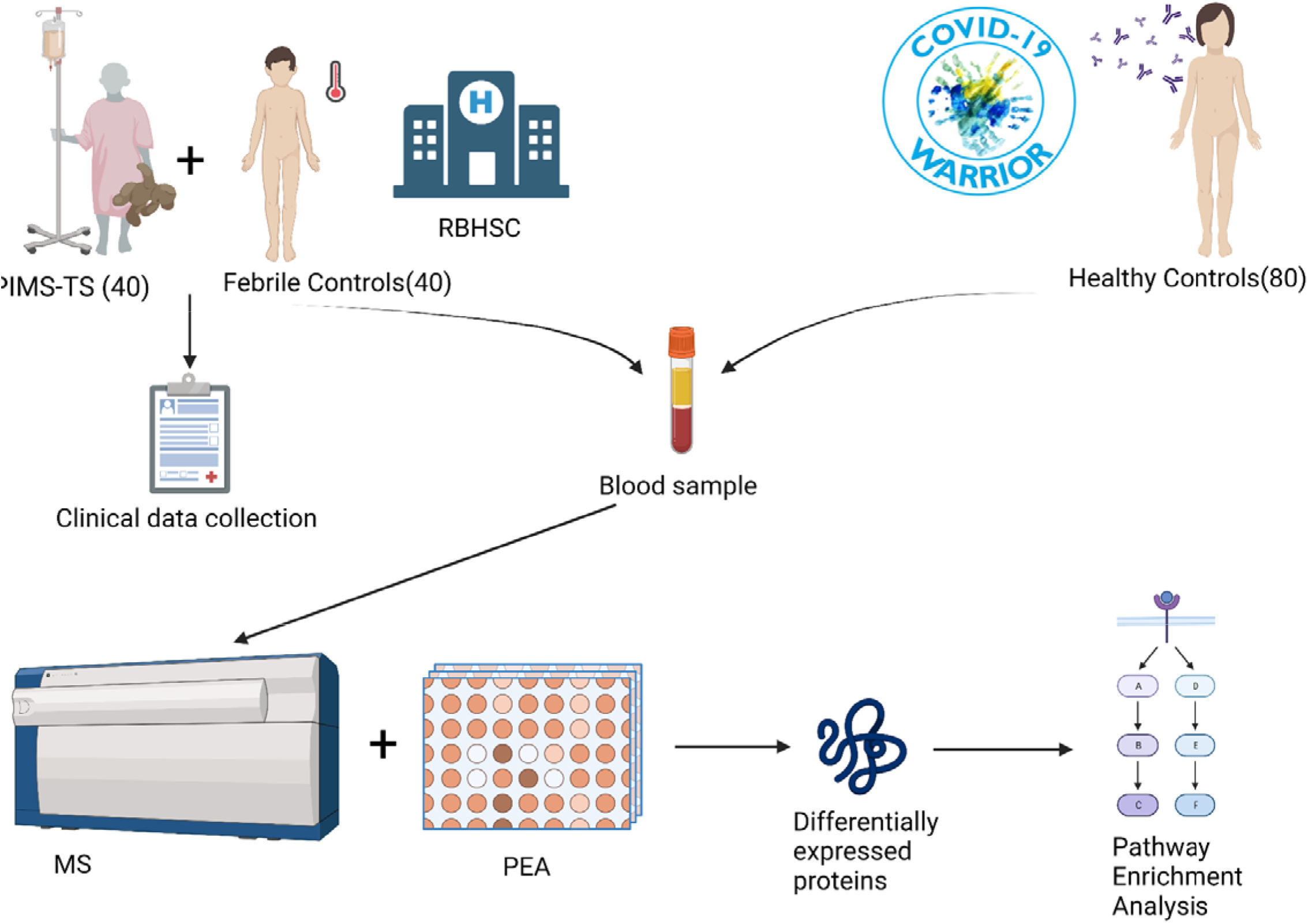
Overview of study design.

### Setting

The Royal Belfast Hospital for Sick Children (RBHSC). The RBHSC is a large tertiary children’s hospital based in the UK. It serves a regional population of 1.9 million people and provides tertiary cardiology care and intensive care. Children with PIMS-TS are typically transferred to either the specialist cardiology ward or the Paediatric Intensive Care Unit (PICU). Eligible participants will be recruited from April 2020 to December 2022.

### Participants

#### PIMS-TS (n=40)

PIMS-TS cases meeting the RCPCH diagnostic criteria are eligible.[1] As such, all PIMS-TS cases must have a persistent fever, evidence of inflammation including elevated C-Reactive Protein (CRP) or inflammatory markers, lymphopenia or neutropenia, and evidence of organ dysfunction. SARS-CoV-2 PCR or serology may be either negative or positive.

#### Febrile Controls

(n=40)

Febrile controls must have a fever on presentation to hospital, and subsequently have a confirmed serious bacterial infection including symptomatic bacteremia (confirmed on blood culture or PCR), meningitis (confirmed on culture or PCR of cerebrospinal fluid), urinary tract infection (confirmed by single growth urine culture and pyuria), pneumonia (confirmed on chest radiograph), bacterial gastroenteritis (confirmed on stool culture) or appendicitis (confirmed on histology). Any children with a known malignancy or have previously received immunotherapy are not eligible as febrile controls. Children with bacterial contaminants on culture will not be eligible for inclusion as febrile controls.

#### Matched Healthy Controls

(n=80)

Healthy children were recruited through the existing Covid Warriors observational cohort study conducted during the same time period, registered at https://www.clinicaltrials.gov on15 April 2020, with identifier NCT04347408.[8] This included children with and without evidence of prior SARS-CoV-2 exposure and seroconversion confirmed on ROCHE nucleocapsid antibody assay.

#### Matching Criteria

Where possible children will be matched based on age and gender. The febrile controls will be matched to the PIMS-TS cases on age and gender. Healthy controls will be chosen from the aforementioned Covid Warriors cohort, with children with evidence of prior SARS-CoV-2 exposure matched to the PIMS-TS patients. Children with no evidence of prior SARS-CoV-2 infection will be matched to the febrile patients. For both patient groups, a one to one ratio between healthy controls and patients will be used. Matching will be done using the ccoptimalmatch package, with controls matched on both age in whole years and gender.[20]

### Recruitment

Potential participants will be approached by a trained researcher and the nature and purpose of the study will be explained. The duration of the study, procedures involved, possibility of discomfort and risks and benefits will be discussed. Parents/Children are free to withdraw from the study at any time, and it will be emphasized that participating in the study will have no bearing on standard of care provided. Consent will also be sought to store samples for use in future.

### Procedures

The only additional procedure is the collection of an additional blood sample (2-5ml). Where possible blood samples will be collected along with routine phlebotomy required for patient care. All phlebotomy will be carried out by experienced paediatric nursing/medical professionals. Blood samples will be collected in an EDTA vacutainer, and Serum separating tube if volume allows. Blood plasma samples will be transported to QUB site by member of study team and immediately processed. A volume of 900ul whole blood will be aliquoted from EDTA tube prior to centrifugation and frozen in liquid nitrogen with 10% DMSO. Multiple aliquots of plasma and serum will be stored at -70C if volume is sufficient. Sample type, volume, date and time of sample collection will be recorded on the RedCap platform.

### Data management and availability

All participant data will be anonymized, with the linkage documentation remaining on site. Anonymized data will be stored securely on RedCap, and General Data Protection Regulation (EU) 2016/679 will be followed. Individuals anonymized data, including raw proteomics results will be available on QUB data repository.

### Ethical approvals

Ethical approval was provided by the London-Chelsea Research ethics committee (REC Reference—20/HRA/1731). Local research governance was provided by the Belfast Health & Social Care Trust Research Governance (Reference 19147TW-SW).

### Funding

This study is funded by the NICHS (code =2021 H3) and the PHA (code = COM/5712/22). The Dr Dinah Kohner studentship also provides fellowship funding.

### Variables

All approaches to potential participants will be recorded in a screening log which will be kept on site. Demographic and clinical data will be recorded for all patients who are recruited This will be completed using a case report form on REDCap, with data collection and entry carried out by an independent paediatric clinician.[21,22] Clinical notes, the Electronic Care Record and Symphony/ED admission notes will be used for data collection. Data will be collected including individuals exposure to SARS-CoV-2, PCR results, symptoms and laboratory values on admission. The case report form to be used for PIMS-TS patients is available in Supplementary info 1.

### Data sources

Clinical data will be recorded from the patient record, using clinical notes, recorded by a clinician. Anonymise data on the healthy controls will be accessed from the Covid Warriors database.

### Bias

To minimise bias the diagnosis of PIMS-TS and serious bacterial infection will be confirmed by independent and experienced Paediatric Intensivists (PC, HNB) and Paediatric Infectious Diseases consultants (HG, SC). They will be blinded to all proteomic analysis.

### Study size

A statistical power analysis was performed for sample size estimation using GPower 3.1.9.7 software. [23] Based on measurement on ntProBNP in PIMS-TS patients and healthy controls by Diorio et al, a minimum of 12 patients per group are required to achieve 90% power to detect difference in means between groups using independent samples two tailed t test at alpha (α) = 0.05 (µ1 = 3.74, n=21, µ2 = 0.115, n=25, σ = 3.62, (f) = 13.12).[24]Based on measurements of ICAM1 in the same group, a minimum of 6 patients per group are required to achieve 90% power at alpha (α) = 0.05 (µ1 = 1.4, n=21, µ2 = 0.279, n=25, σ = 1.12, (f) = 3.32). Thus, with n=20 patients per group for the proposed study we have sufficient sample numbers to identify biomarkers of statistical significance.

### Statistical methods

Quantitative Mass spectrometry data will be exported from DIA-NN to excel files. Data will be normalized and filtered to proteins present in >70% of biological samples. Proteins with >20% Coefficient Variation in technical replicate samples will be filtered. The filtered, normalized data for biological samples will be used for Principal component analyses (PCA) and hierarchical cluster analyses, to easily visualize the high dimensional data and explore sample groupings.

Data from PEA’s will be provided pre-processed as NPX values. Proteins with intra-assay CV’s >15% or inter-assay CV’s > 30% will be excluded. NPX values from Immune response and Inflammation panels will be merged and analysed together, while NPX values from the cardiometabolic panel will be analysed separately. Again, PCA and unsupervised clustering will be used to visualize sample groupings.

Separate statistical approaches for identifying candidate biomarkers and for identifying proteins involved in pathways underlying PIMS-TS pathogenesis. An unpaired t test with false discovery rate correction will be used to compare means of proteins between the PIMS-TS and febrile controls groups. Those proteins with FDR adjusted p value less than 0.05 and a log2 fold change of at least 1 will be identified as significantly differentially expressed. Significant proteins will be assessed for relationship with age by linear regression.

### Pathway analysis

An alternative approach will be used to identify proteins to be used for pathway analysis. Uncorrected t tests between PIMS-TS and healthy control groups, PIMS-TS and febrile controls and febrile controls and healthy controls will be performed. From these tests proteins which are significantly different between PIMS-TS and both febrile and healthy control groups will be identified and used for pathway analysis. This will be performed using the pathfindr package with a variety of gene sets such as “REACTOME” and “KEGG”.[25] Protein ID’s and log2 fold change between PIMS-TS and febrile controls will be uploaded. Significantly enriched terms will be identified.

Subgroup analyses of the PIMS-TS and febrile cohorts will also be performed. Correlation between clinical severity and levels of significantly different proteins will be assessed. An uncorrected T test will be used to compare those PIMS-TS patients who required ICU admission to those who did not.

### Proximity extension assay

Plasma samples from PIMS-TS, febrile and healthy controls will be randomized in a 96 well format. A total of 537 unique proteins will be measured using the Inflammation, Immune Response Target 96 and Cardiometabolic Explore proximity extension assay panels provided by Olink. [26,27] Each panel consists of a plate with each well containing 96 pairs of oligonucleotide tagged analyte specific antibodies. Four internal controls and six sample controls are ran to allow for quality control of inter plate variation. The CT values from the qPCR reaction are converted to Normalised Protein Expression (NPX) values, which is an arbitrary unit on log2 scale. The calculation of NPX values differs between the Target 96 and Explore panel, and cannot be directly compared.

### Mass Spectrometry sample preparation

Plasma samples will be used for mass spectrometry. All samples will be prepared in a 96 well format which allows for high throughput preparation of large sample numbers while minimizing technical variation. A 96 deep well filter plate format will be used for plasma depletion of most abundant proteins. This improves the detection of lower abundant proteins, which can be difficult to detect in undepleted plasma.[28,29] Order of samples on plate will be randomized using Excel random number generator, and samples will be recoded by an independent lab member. Once depleted, samples can be frozen and stored, or can be used for tryptic digestion in a 96 well plate format. A sample pool of depleted samples will be made and digested along with biological samples. Digested samples can be frozen or desalted using a modification of stage tipping which allows 96 samples to be desalted at once. [30,31]A pool of digested samples will be used for fractionation.

### Depletion of most abundant plasma proteins

High Select TOP14 resin is homogenized on rollers. 300ul of resin aliquoted into bottom of wells in 96 well plate. 10ul of each plasma sample is diluted 1:10 in 0.05M ABC. 10ul of this diluted plasma is added to wells. Plate is covered with parafilm and incubated for 20 minutes on plate shaker at 150 oscillations/min. Place 96 well collection plate beneath the depletion plate and place in a swinging bucket rotor centrifuge. Centrifuge for 2 minutes and 100xG. Remove plates and separate. Depleted sample will have flowed through to collection plate. Depleted sample plate will be dried down in vacuum concentrator. Resuspend pellets in 100ul 0.05M ABC. Add 400ul cold acetone and keep for 16 hours at -20 ºC. Spin at 15000xg for 15minutes. Remove supernatant using pipette with care. Dry sample using vacuum concentrator to eliminate acetone residue.

### Sample Digest

Resuspend pellets in 100ul 8M Urea, 0.1M Tris-HCL pH8.0. Measure sample protein concentration using BCA. Aliquot 2ul from each sample to new cryovial to create a depleted sample pool. Randomize sample order and include a well with a sample pool on each well of 96 well plate layout.Aliquot 50ug proteins equivalent of sample volume to new 96 well plate and add 5ul 100mM Dithiothreitol(DTT). Incubate at 27 Celsius for 1 hr. Spin at room temperature. Add 5ul 140mM iodoacetamide and incubate at room temperature for 30 minutes in the dark. Add 2ul 100mM DTT. Add 108ul 0.05M ABC to dilute urea. Add 5ul Trypsin, (0.2ug/ul) and incubate for 16 hours on thermomixer in 37 Celsius incubator.Add 2ul 100% Trifluoroacetic acid (TFA) to a 1% final concentration to quench the digest.

### Sample desalting

Dry samples using vacuum concentrator. Resuspend samples in 20ul 1% TFA and measure sample peptide concentration using Peptide Quant kit. Prepare necessary number of stage tips using 200ul pipette tips and c18 extraction discs. All centrifugation steps are done for 4 minutes at 4700rpm. Condition stage tip by adding 50ul 50% Acetonitrile, 0.1% TFA and centrifuge. Add 50ul 1%TFA and centrifuge. Aliquot volume equivalent to 8ug peptide into stage tip and centrifuge. Add 50ul 1%TFA, centrifuge and repeat. Move stage tips to individual eppendorfs. Add 25ul ACN 50%, 0.1% TFA centrifuge and repeat. Remove stage tip from Eppendorf. Dry down flow through in vacuum concentrator and resuspend in 20ul Buffer A.

### Mass Spectrometry

All samples will be run on the same session. Digested samples will undergo unbiased, deep proteomic analysis on a timsTof Pro mass spectrometer (Bruker Daltonics) connected to a Evosep One liquid chromatography system(EvoSep BioSystems). The peptides will be separated on a reversed-phase C18 Endurance column using preset 30 SPD method. Analysis of high pH-reversed phase fractionated sample pools will be performed in data dependent acquisition mode (dda) to generate spectral library data. Individual samples will be analysed in data independent acquisition mode (dia). All data will be acquired with the instrument operating in trapped ion mobility spectrometry (TIMS) mode. Trapped ions will be selected for ms/ms using parallel accumulation serial fragmentation (PASEF). As a quality control, pooled sample digests will be analysed at regular intervals throughout the run. Raw data files will be processed through DIA-NN software for spectral library building, protein identification and quantification.[32]

### Validation of candidate diagnostic biomarkers

The top 5 proteins with largest fold change in the identified differentially expressed proteins between PIMS-TS and Febrile controls will be measured in our cohort’s plasma samples using Enxyme-linked Immunosorbent assay. If possible, these proteins will be verified in an independently recruited cohort of PIMS-TS patients. Failing this, there are publicly available datasets of PIMS-TS proteomes which may be accessed for in silico verification. [33,34]

### In vitro modelling of PIMS-TS

The well documented endothelial involvement in PIMS-TS will be investigated in an invitro model, using a Human Umbilical Vein Endothelial cell (HUVEC) line. [24,35] Patient sera will be used to treat HUVEC’s for a range of durations, at a range of concentrations from 0 to 15% patient sera. PIMS-TS, Healthy Control and febrile control sera will be used. Western blot and quantitative PCR will be used to quantify changes in protein and genes of interest abundance. In addition, based on pathway enrichment analysis results an in vitro model of PIMS-TS will be designed with a suitable cell line to investigate the top enriched pathway further.

## Discussion

This study will be the first to fully characterize the proteome of PIMS-TS by combining both Mass Spectrometry and PEA technology. It will also be the first to compare the proteome of children with PIMS-TS to that of healthy children who have been exposed to SARS-CoV-2. The comparison between PIMS-TS and febrile controls will allow potential diagnostic biomarkers to be identified. The differences in proteome of healthy children following SARS-CoV-2 infection and those who progress to PIMS-TS may allow underlying disease driving mechanisms to be identified. This antibody positive healthy cohort is a particular advantage of this study, which has not been available to other studies investigating the PIMS-TS proteome.[24,33,36,37]

Limitations-This study may be limited by sample number due to the rarity of PIMS-TS. The PIMS-TS population recruited may not be similar to those in other countries.

## Data Availability

All data produced in the present study are available upon reasonable request to the authors.

## Notes

### Competing Interest Statement

The authors have declared no competing interest.

### Clinical Protocols

https://bmjopen.bmj.com/content/10/11/e041661

https://adc.bmj.com/content/106/7/680

### Funding Statement

This work was supported by HSC R&D Division, Public Health Agency
Ref: COM/5596/20, Northern Ireland Chest Heart and Stroke 2021_H03, and the Dr Dinah Kohner Studentship. These funding sources had no role in the design of this study and
will not have any role during its execution, analyses, interpretation of the data, or
decision to submit result.

### Author Declarations

Ethical approval to conduct this study was provided by Belfast Health & Social Care Trust Research Governance (Reference 19147TW SW) and the London Chelsea Research ethics committee (REC Reference 20/HRA/1731).

