## Supplementary information 1. for "Study protocol: Medium throughput, deep proteomic characterization of children with PIMS-TS, and identification of candidate diagnostic biomarkers"

### PIMS clinical characteristics

| | |
|---|---|
| Study ID | __________________________________ |
| RAPID-19 ID | __________________________________ |
| Sex | ◯ Female<br>◯ Male |
| District General Hospital transfer | ◯ No<br>◯ Yes |
| Past Medical History | ◯ No<br>◯ Yes |
| PMH : ex-preterm(< 36 weeks) | ◯ No<br>◯ Yes |
| pmh: cardiac | ◯ No<br>◯ Yes |
| PMH: resp | ◯ No<br>◯ Yes |
| PMH: Neuro | ◯ No<br>◯ Yes |
| PMH:GI | ◯ No<br>◯ Yes |
| PMH: allergy/atophy | ◯ No<br>◯ Yes |
| PMH:Other | ◯ No<br>◯ Yes |
| Other PMH specified | __________________________________________ |
| Covid-19 Vaccine received | ◯ No<br>◯ Yes |
| Symptomatic COVID-19 within 2 months of onset | ◯ No<br>◯ Yes |
| Regular medication inc antibiotics | ◯ No<br>◯ Yes |
| Specify medication | __________________________________________ |

Duration of fever (>38oC) prior to hosp (days) __________________________________

#### PIMS Symptoms

| | No | Yes |
|---|---|---|
| Cough | ◯ | ◯ |
| Sore Throat | ◯ | ◯ |
| Runny Nose | ◯ | ◯ |
| Wheeze | ◯ | ◯ |
| SOB | ◯ | ◯ |
| Vomiting | ◯ | ◯ |
| Abdo Pain | ◯ | ◯ |
| Diarrhoea | ◯ | ◯ |
| Muscle aches/Arthritis | ◯ | ◯ |
| Headache /Meningism | ◯ | ◯ |
| Confusion/Drowsiness | ◯ | ◯ |
| Collapse/Syncope | ◯ | ◯ |
| Conjunctivitis | ◯ | ◯ |
| Cervical lymphadenopathy | ◯ | ◯ |
| Rash | ◯ | ◯ |
| Lip/Mucosal changes | ◯ | ◯ |
| Extremity changes | ◯ | ◯ |

Admission:HR __________________________________

Admission: CRT>2s
◯ No
◯ Yes

Admission:Systolic BP __________________________________

Admission: Concern for Shock
◯ No
◯ Yes

Admission: SaO2 in RA __________________________________

SaO2< 90%
◯ No
◯ Yes

Admission: GCS Abnormal
◯ No
◯ Yes

Initial ECG reported as abnormal (other than sinus tachy)
◯ No
◯ Yes

#### Initial Blood work

| | |
|---|---|
| Initial CRP (mg/L) | __________________ |
| Initial Hb (g/dL) | __________________ |
| Initial WBC (x10^9/L) | __________________ |
| Initial Neutrophil count( x10^9/L) | __________________ |
| Initial Lymphocyte count (x10^9/L) | __________________ |
| Initial PLT(x10^9/L) | __________________ |
| Initial Sodium (umol/L) | __________________ |
| Evidence of Acute Renal Failure | ◯ No<br>◯ Yes |
| Initial APPT (Sec) | __________________ |
| Initial Fibrin (g/L) | __________________ |
| Initial D-Dimer(mg/L) | __________________ |
| Initial Troponin (ng/L) | __________________ |
| Initial ProBNP (ng/L) | __________________ |
| Initial CK (u/L) | __________________ |
| Initial LDH (U/L) | __________________ |

#### Subsequent results

| | |
|---|---|
| Lowest Hb (g/dL) | ______________________ |
| highest neut (x10^9/L) | ______________________ |
| lowest lymph (x10^9/L) | ______________________ |
| lowest PLT (x10^9/L) | ______________________ |
| highest CRP (mg/L) | ______________________ |
| Duration CRP above 100 (days) | ______________________ |
| Highest ALT (IU/L) | ______________________ |
| Lowest Alb (g/L) | ______________________ |
| highest trop (ng/L) | ______________________ |
| Highest proBNP (ng/L) | ______________________ |
| highest ferritin (ug/L) | ______________________ |
| highest D-dimer (mg/L) | ______________________ |
| Positive co-infection test for other resp virus | ◯ No<br>◯ Yes |
| Specify co-infection(viral) | ______________________ |

#### Treatment

| | No | Yes |
|---|---|---|
| IVIG | ◯ | ◯ |
| Repeat IVIG | ◯ | ◯ |
| Steroids < =3 days | ◯ | ◯ |
| Steroids> 3 days | ◯ | ◯ |

| | | |
|---|---|---|
| Immunomodulator | ◯ | ◯ |
| Aspirin | ◯ | ◯ |

Immunomodulator specify ______________________

#### Initial outcome measures

ICU Admission
◯ Yes
◯ No

Inotroph use
◯ Yes
◯ No

duration of hospital stay (days) ______________________

Initial ECHO abnormal coronary arteries
◯ Yes
◯ No

initial Ejection fraction abnormal?
◯ Yes
◯ No

Long term outcome measure: Follow-up ECHO abnormal
◯ Yes
◯ No

Specify echo abnormalities ______________________
